# Cardiometabolic and Psychobehavioral Phenotypes Define Cardiovascular Risk Heterogeneity in Rheumatoid Arthritis

**DOI:** 10.64898/2026.09.17.26363297

**Authors:** Shah Tanzen Jahan, Anicha Akter, Sojib Hossain, Md Selim Reza, Md. Mijanur Rahman, Md. Mahmudul Alam, Md. Monimul Huq

## Abstract

Cardiovascular disease (CVD) is a major cause of morbidity and premature mortality in people with rheumatoid arthritis (RA). Whether multidimensional cardiometabolic, inflammatory, renal, behavioral, and psychosocial factors define distinct cardiovascular risk phenotypes in RA remains unclear. This study aimed to identify data-driven phenotypes and evaluate their associations with prevalent CVD.

We analyzed 3,252 adults aged≥20 years with self-reported RA from the 2005–2018 National Health and Nutrition Examination Survey (NHANES). Unsupervised partitioning around medoids (PAM) clustering using Gower distance identified distinct data-driven cardiovascular-risk profiles. Cluster validity was assessed using silhouette analysis and internal train–test validation. Survey-weighted binary logistic regression evaluated associations between phenotype membership and prevalent CVD.

Six clinically interpretable data-driven phenotypes were identified with varying CVD prevalence: Severe Metabolic–Diabetic (40.3%), Aging Diabetic–Hypertensive (34.9%), Non-Diabetic Intermediate (20.2%), Mild Metabolic (14.8%), Cardiometabolic (10.4%), and Smoking-Predominant (19.4%). Compared with the Cardiometabolic phenotype, the Severe Metabolic–Diabetic phenotype exhibited the highest odds of prevalent CVD (aOR 4.11, 95%CI:2.56–6.59), followed by the Aging Diabetic–Hypertensive phenotype (aOR 3.38, 95%CI:2.22–5.15). Higher odds of prevalent CVD were also observed in the Smoking-Predominant (aOR 1.99, 95% CI: 1.26–3.13) and Non-Diabetic Intermediate (aOR 1.77, 95% CI: 1.10–2.84) phenotypes. The six-cluster solution demonstrated moderate separation and strong internal correspondence across training and testing sets, while the phenotype-informed model showed moderate discrimination for prevalent CVD (AUC = 0.72).

Among adults with rheumatoid arthritis (RA), distinct data-driven phenotypes were identified based on combined patterns of metabolic, inflammatory, renal, behavioral, and psychosocial factors, with differing burdens of prevalent cardiovascular disease (CVD). Phenotype-based approaches may provide a comprehensive framework for characterizing multidimensional cardiovascular risk in RA.

## Introduction

Rheumatoid arthritis (RA) is a chronic systemic autoimmune disorder that primarily affects the synovial joints and multiple organ systems[1]. Its global prevalence is approximately 0.25% to 1% and projected to reach approximately 31.7 million by 2050 [2], [3]. Patients with RA have a 1.5–2 fold higher risk of cardiovascular disease (CVD) compared to those without RA [4]. This excess risk has been linked to chronic inflammation, endothelial dysfunction, and metabolic alterations that are not fully captured by traditional risk models[5], [6]. Accordingly, the European League Against Rheumatism (EULAR) has formally recognized RA as an independent cardiovascular risk (CVR) condition and recommends its management as an integral component of RA care[7]. Despite this recognition, the translation of these recommendations into individualized, precision-oriented risk stratification strategies remains insufficient.

CVD risk assessment in RA has largely focused on individual biomarkers or risk factors in isolation[8]. Such approaches are insufficient to capture the multidimensional nature of CVR in this population. Cardiometabolic, behavioral, and psychosocial factors including depression, physical inactivity, and smoking interact in ways not adequately represented in traditional risk models[9][10]. In addition, standard CVR calculators perform poorly in RA, underestimating true risk[11]. Beyond these limitations, RA is a heterogeneous condition characterized by substantial variation in metabolic profile, inflammatory burden, comorbidity patterns, and health behaviors[12]. Treating RA as a homogeneous entity may obscure clinically meaningful subgroups and obscure clinically meaningful subgroups with differing CVR profiles[13].

Existing evidence has demonstrated heterogeneity in CVR among patients with RA, often through subgroup analyses based on demographic, clinical, or inflammatory characteristics[14][15], [16]. However, these approaches largely rely on investigator-defined categories based on a limited number of variables. Such frameworks may not fully capture the complex, multidimensional interactions associated with CVR in RA. Data-driven methods that allow phenotypes to emerge empirically from integrated biological, behavioral, and psychosocial factors may therefore provide a more comprehensive characterization of risk.

The present study employed a clustering algorithm to simultaneously integrate cardiometabolic, inflammatory, renal, behavioral, and psychosocial variables, rather than stratifying patients by individual variables or clinical categories. This allows data-driven phenotypic subgroups to emerge empirically from the data structure without a priori assumptions[15]. This approach moves beyond the binary or tertile-based subgroup contrasts of prior work[17], [18] and captures the multivariate co-occurrence of risk factors observed in patients with RA[19]. Critically, the simultaneous inclusion of depression and physical inactivity alongside cardiometabolic and inflammatory markers addresses a dimension of integrated psychobehavioral risk that prior RA cardiovascular stratification models have not systematically incorporated. this study complements prior approaches by providing an empirically grounded, multidimensional characterization of cardiovascular-risk profiles in adults with RA that may support further evaluation of phenotype-based approaches to CVR assessment.

## Materials and Methods

### Data source and study population

This cross-sectional study used data from seven NHANES cycles spanning 2005–2018. RA was identified using self-reported physician diagnosis. Participants were classified as having RA if they answered “yes” to the NHANES Medical Conditions Questionnaire (MCQ) item asking whether a doctor or other health professional had ever informed them that they had RA. A total of 3,496 participants met this definition of RA.

The final analytic sample was refined using the prespecified eligibility and exclusion criteria (Figure 1). Participants aged ≥20 years were included, consistent with established NHANES/NCHS conventions for adult examination-based analyses. Participants were excluded if they were pregnant; had zero or missing Mobile Examination Center (MEC) examination weights; had missing data for the primary outcome; had missing age, sex, or survey design-related information; or had extreme values of BMI (>60 kg/m²), systolic blood pressure (SBP >220 mmHg), triglycerides (>1,000 mg/dL), NLR (>15), or eGFR (<10 mL/min/1.73 m²). Finally, 3,252 adults with RA were included in the analytic sample.

**Figure 1.**
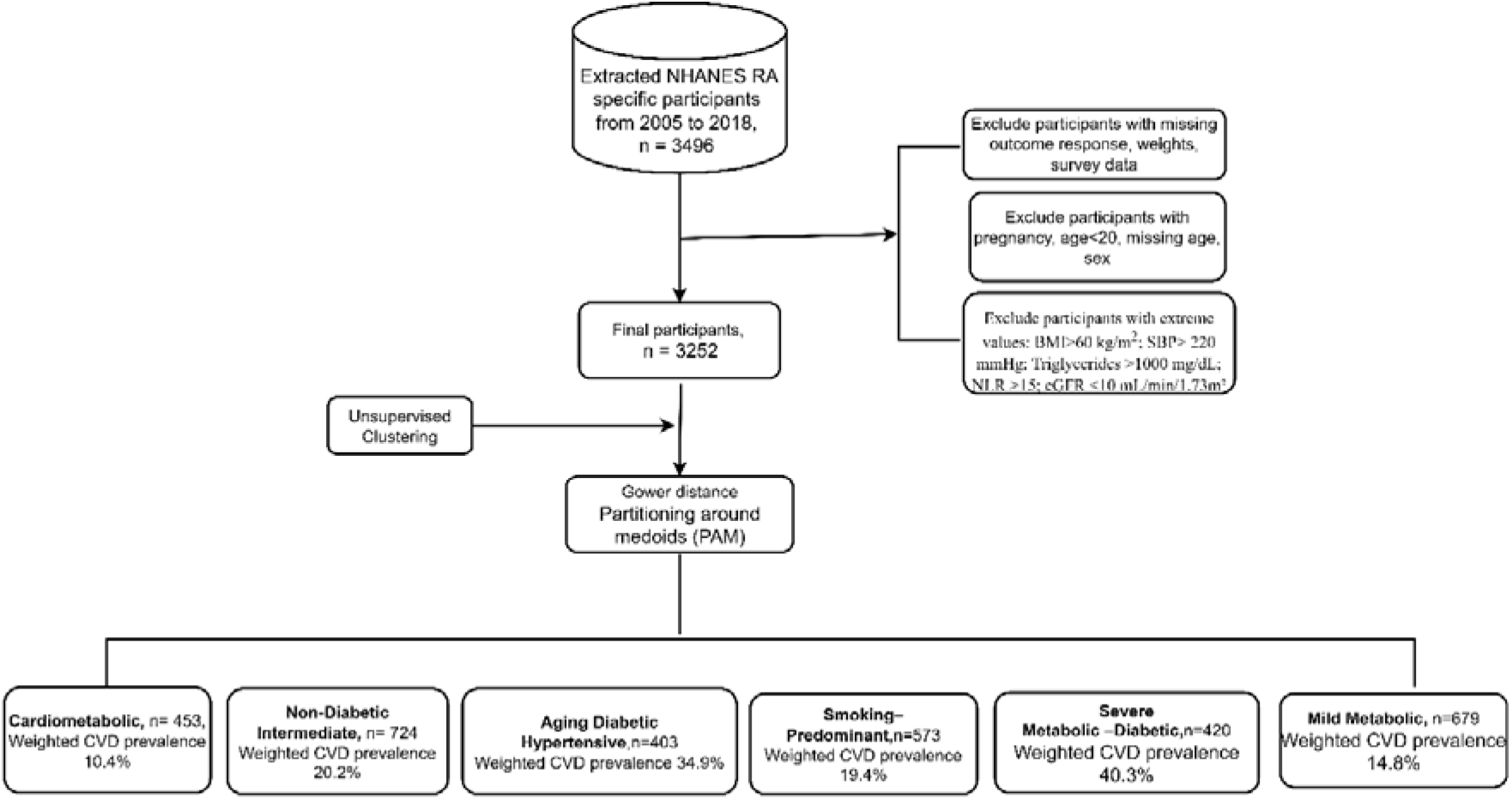
Participant selection and clustering workflow. Flow diagram illustrating the selection of the analytic RA cohort from NHANES 2005–2018, application of the prespecified exclusion criteria, clustering of the entire analytic cohort using Gower distance and partitioning around medoids (PAM), identification of six data-driven phenotypes, and subsequent evaluation of CVD prevalence.

### Assessment of cardiovascular disease

CVD status was ascertained using data from the medical condition questionnaire (MCQ) component. This component captures self-reported physician diagnoses obtained through standardized in-person interviews. Participants were classified as having CVD if they reported a prior physician diagnosis of any of the following conditions: angina, coronary heart disease, myocardial infarction, congestive heart failure, or stroke. A composite CVD variable was constructed to represent the presence of one or more of these conditions.

### Covariates measures

In this study, we considered age, and ratio of family income to poverty (PIR) as continuous variable, Sex was categorized as male or female, race had five categories as Mexican American, other Hispanic, Non-Hispanic White, Non-Hispanic Black, other race including multiracial, smoking status were classified as smoker (currently, former) and non-smoke (never), physical activity had yes and no categories.

Body Mass Index (BMI, kg/m²) was treated as a continuous variable, whereas triglyceride levels were log-transformed because of their right-skewed distribution. Systemic inflammation was assessed using the neutrophil-to-lymphocyte ratio (NLR), which was also log-transformed because of its right-skewed distribution (see Supplementary Figure S6). Depression was calculated through a total severity score by summing the responses from the PHQ-9 modules (variables DPQ010 through DPQ090), which assess the frequency of depressive symptoms over the past two weeks. Participants with a PHQ-9 score ≥10 were categorized as depressed, whereas those with a score <10 were categorized as not depressed[20].

In our study, diabetes was classified as having diabetes (yes/no) if participants reported a previous clinical diagnosis or had a glycated hemoglobin (HbA1c) level ≥6.5%. HbA1c provides an established biochemical criterion for diabetes without requiring fasting, allowing diabetes status to be assessed consistently across the analytic sample. Fasting plasma glucose was not incorporated because its use would require restriction to the NHANES fasting subsample, potentially reducing the available sample size and comparability across participants. Hypertension was defined based on the average of three systolic and diastolic blood pressure measurements. Mean systolic blood pressure (SBP) and mean diastolic blood pressure (DBP) were calculated, and a binary indicator (yes) was assigned if mean SBP ≥ 130 mmHg, mean DBP ≥ 80 mmHg, or if the participant reports a prior diagnosis of high blood pressure (BPQ020). Estimated glomerular filtration rate (eGFR) was calculated via the simplified Modification of Diet in Renal Disease (MDRD) equation[21]. Details on the measurement of covariates can be found on the NHANES website (https://www.cdc.gov/nchs/nhanes/index.htm). Variables were selected based on clinical relevance and prior literature linking these factors to cardiovascular risk in RA.

Age, sex, race, and PIR were not included in the clustering procedure. Instead, these variables were treated as covariates and were incorporated into survey-weighted binary logistic regression models to evaluate the independent association between phenotype membership and CVD.

### Statistical Analysis

All statistical analyses were conducted in accordance with National Center for Health Statistics (NCHS) guidelines(https://wwwn.cdc.gov/nchs/nhanes/tutorials/default.aspx), accounting for the complex multistage, stratified probability sampling design where appropriate. The unsupervised clustering analysis was performed on the analytic participant-level data using the prespecified clustering variables, without incorporating survey weights, strata, or primary sampling units into the Gower distance or partitioning around medoids (PAM) procedure. Survey weights and the NHANES complex survey design were subsequently incorporated into descriptive and regression analyses to obtain population-level estimates. The Mobile Examination Center (MEC) examination weight (WTMEC2YR), strata (SDMVSTRA), and primary sampling units (SDMVPSU) were used. Because seven consecutive 2-year NHANES cycles from 2005–2006 through 2017–2018 were pooled, the 2-year MEC examination weight was divided by seven to construct the combined examination weight (WTMEC2YR/7). The combined examination weight, strata, and primary sampling units were incorporated into the survey design for all survey-weighted analyses.

Missing data were addressed using multiple imputation by chained equations (MICE)[22]. All variables included in the analysis had less than 10% missingness. Convergence of the imputation process was assessed through trace plots, density and strip plots (see Supplementary figure S1).

Descriptive statistics were used to summarize sociodemographic and clinical characteristics. To understand the association between CVD and covariates such as age, sex, BMI, race, blood pressure etc. we performed weighted univariate test such as Design-based Kruskal Wallis test for continuous variables, and Rao & Scott adjustment for categorical variables.

### Cluster Analysis

The primary exposure of interest in this study was membership in data-driven cardiometabolic and psychobehavioral phenotypes. To characterize the determinants of CVR in RA, ten variables were selected a priori for the clustering analysis based on biological plausibility and evidence from previous literature[7][23]. These variables encompassed five complementary domains: (1) metabolic factors, including BMI, triglycerides, high-density lipoprotein cholesterol (HDL-C), diabetes mellitus, and hypertension; (2) inflammatory burden, represented by the NLR; (3) renal function, assessed using eGFR; (4) behavioral factors, including smoking status and physical activity; and (5) psychosocial status, represented by depression. These variables were entered into the clustering algorithm and served as phenotype-defining indicators rather than outcome variables or adjustment covariates.

Unsupervised clustering was performed to identify distinct cardiometabolic and psychobehavioral phenotypes among participants with RA [22]. Clustering variables were selected to represent multidimensional determinants of CVR. Gower distance was used to compute pairwise dissimilarities between individuals. Partitioning around medoids (PAM) clustering was then applied to the resulting distance matrix. This method is suitable for mixed data structures[24]. The optimal number of clusters was determined by evaluating average silhouette width across candidate solutions (k = 2–7). Cluster structure was further visualized using t-distributed stochastic neighbor embedding (t-SNE) and standardized cluster-specific clinical profiles. Final cluster assignments were retained for downstream analyses. Phenotype labels were assigned post hoc based on dominant clinical characteristics from weighted descriptive profiles. These labels were used to facilitate clinical interpretation and did not influence the clustering process. Weighted mean and standard error (S.E) were reported in the result for continuous variables and percentage for categorical variables.

Internal validation was performed using a fixed-seed 70:30 train–test split. The complete clustering pipeline was independently repeated in both cohorts using identical variables. Cluster reproducibility was assessed by comparing average silhouette width, Davies–Bouldin, Dunn, and Calinski–Harabasz indices between datasets.[25]. Phenotype similarity between corresponding train and test clusters was quantified using cosine similarity of standardized phenotype profiles. Visual reproducibility was further evaluated using t-SNE projections (see Supplementary S4) and standardized heatmaps (see Supplementary S5).

Binary survey-weighted logistic regression models were used to evaluate the association between cluster membership and CVD. These models were adjusted for sociodemographic factors, including age, sex, race, and the PIR. Multicollinearity among covariates was evaluated using the Variance Inflation Factor (VIF), with a threshold of < 5 (see supplementary table S2).

Nonlinear associations between estimated glomerular filtration rate (eGFR) and prevalent CVD were evaluated using restricted cubic splines (RCS) with three knots. The overall association and nonlinear component were assessed using Wald tests. Model discrimination for prevalent CVD was evaluated using the area under the receiver operating characteristic curve (AUC). All the analysis were performed using the R software version R 4.6.0.

## Results

### Baseline Characteristics of the Study Population

The study included 3,252 participants, representing an estimated U.S. population of 20,459,215 adults. The prevalence of CVD was 20.62% among participants with RA. Table 1 summarizes the weighted baseline demographic, clinical, and lifestyle characteristics of the participants stratified by CVD status. Unweighted baseline characteristics are provided in supplementary Table S1.

**Table 1:**
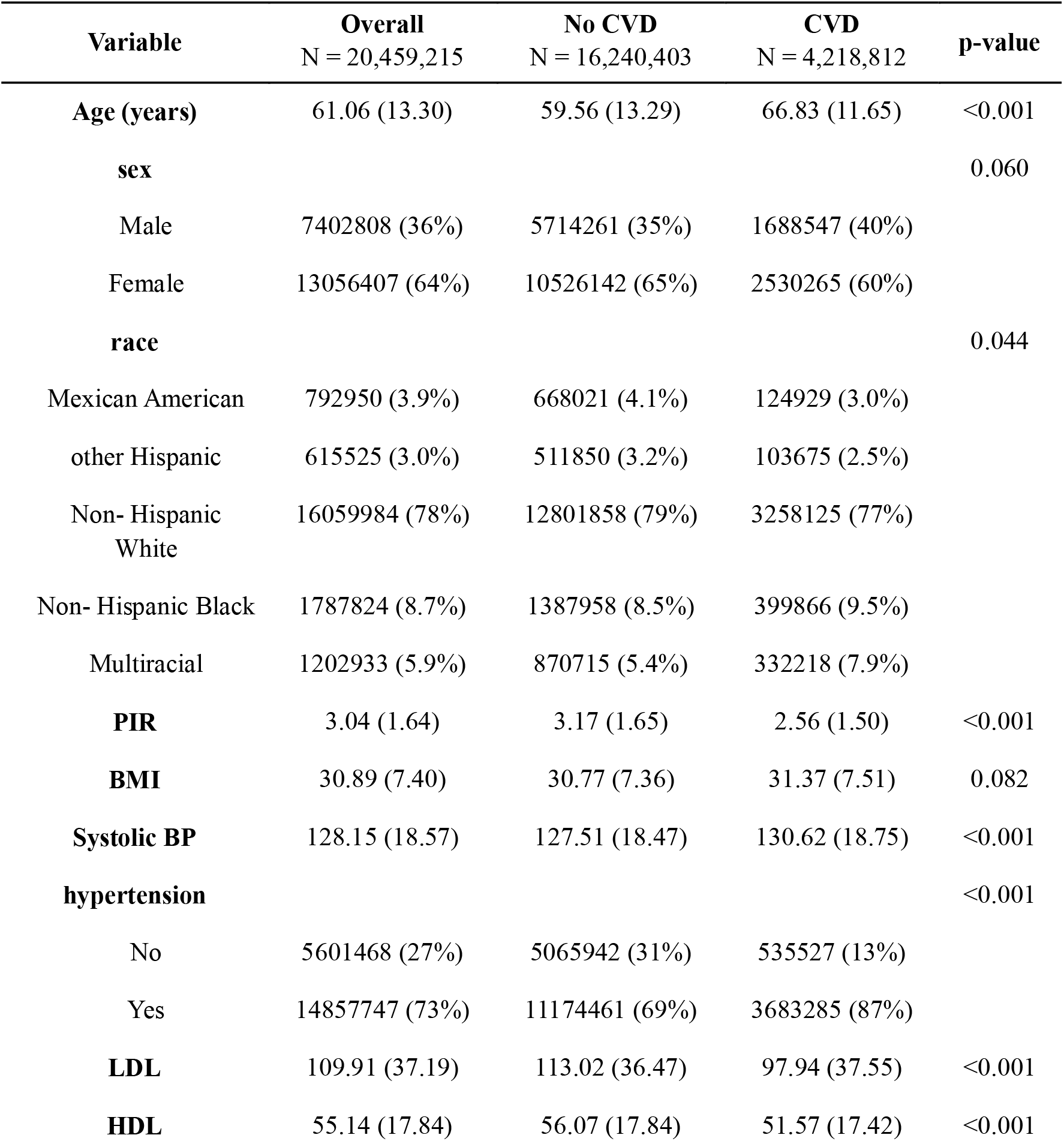
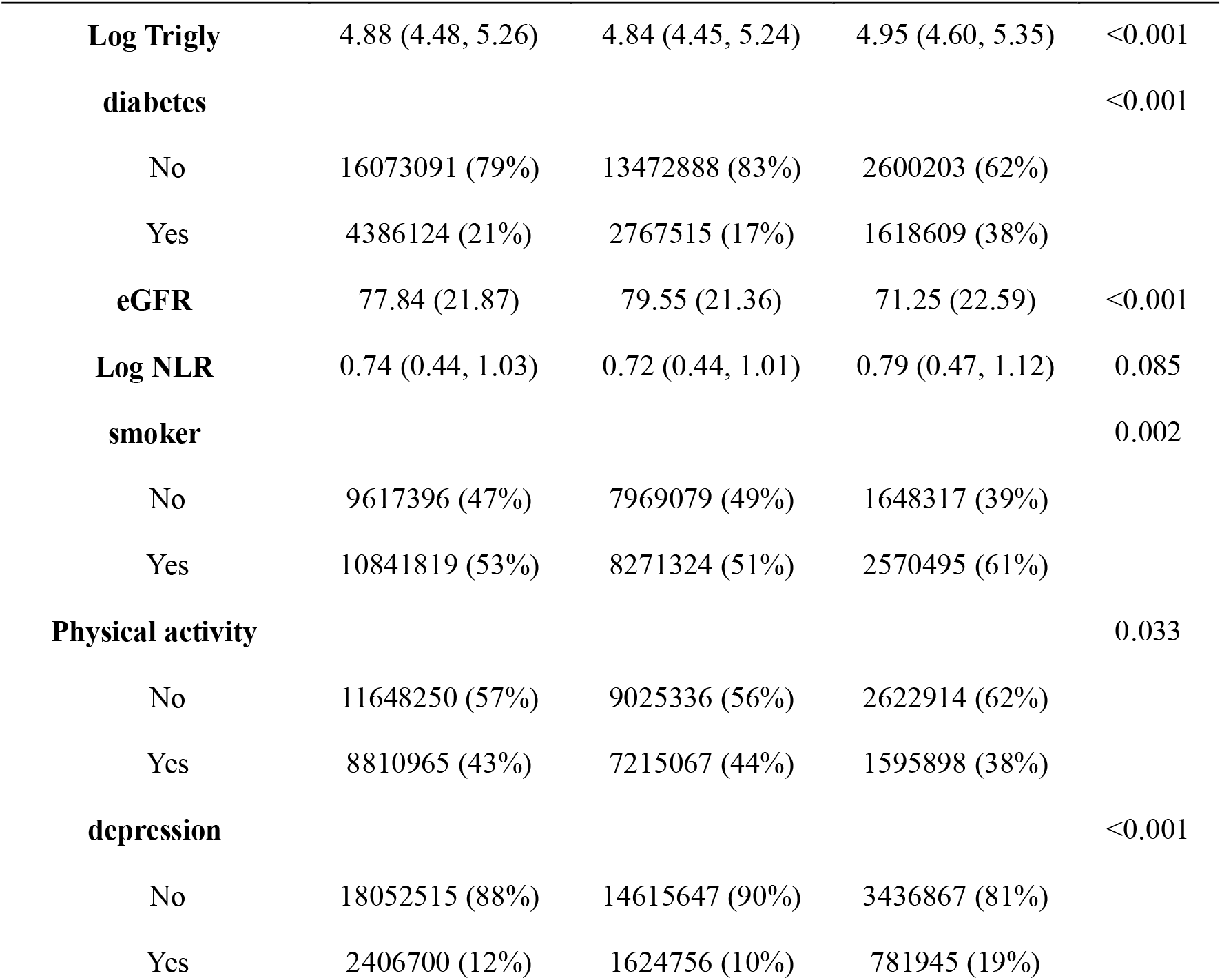
Weighted Baseline Demographics, Cardiometabolic Risk Factors, and Psychobehavioral Characteristics of RA Population. Values are presented as weighted continuous means (SD), medians (IQR), or categorical frequencies (%). The total sample represents a weighted target population of N = 20459215 individuals stratified by CVD status. Continuous variables were evaluated using design-adjusted survey Wald tests, or the design-based Kruskal-Wallis test. Categorical variables were evaluated using Pearson’s Chi-square (χ^2^) test with the Rao & Scott adjustment.

| Variable | Overall<br>N = 20,459,215 | No CVD<br>N = 16,240,403 | CVD<br>N = 4,218,812 | p-value |
| --- | --- | --- | --- | --- |
| **Age (years)** | 61.06 (13.30) | 59.56 (13.29) | 66.83 (11.65) | <0.001 |
| **sex** |  |  |  | 0.060 |
| Male | 7402808 (36%) | 5714261 (35%) | 1688547 (40%) |  |
| Female | 13056407 (64%) | 10526142 (65%) | 2530265 (60%) |  |
| **race** |  |  |  | 0.044 |
| Mexican American | 792950 (3.9%) | 668021 (4.1%) | 124929 (3.0%) |  |
| other Hispanic | 615525 (3.0%) | 511850 (3.2%) | 103675 (2.5%) |  |
| Non- Hispanic<br>White | 16059984 (78%) | 12801858 (79%) | 3258125 (77%) |  |
| Non- Hispanic Black | 1787824 (8.7%) | 1387958 (8.5%) | 399866 (9.5%) |  |
| Multiracial | 1202933 (5.9%) | 870715 (5.4%) | 332218 (7.9%) |  |
| **PIR** | 3.04 (1.64) | 3.17 (1.65) | 2.56 (1.50) | <0.001 |
| **BMI** | 30.89 (7.40) | 30.77 (7.36) | 31.37 (7.51) | 0.082 |
| **Systolic BP** | 128.15 (18.57) | 127.51 (18.47) | 130.62 (18.75) | <0.001 |
| **hypertension** |  |  |  | <0.001 |
| No | 5601468 (27%) | 5065942 (31%) | 535527 (13%) |  |
| Yes | 14857747 (73%) | 11174461 (69%) | 3683285 (87%) |  |
| **LDL** | 109.91 (37.19) | 113.02 (36.47) | 97.94 (37.55) | <0.001 |
| **HDL** | 55.14 (17.84) | 56.07 (17.84) | 51.57 (17.42) | <0.001 |

| **Variable** | **Overall**<br>N = 20,459,215 | **No CVD**<br>N = 16,240,403 | **CVD**<br>N = 4,218,812 | **p-value** |
| --- | --- | --- | --- | --- |
| **Log Trigly** | 4.88 (4.48, 5.26) | 4.84 (4.45, 5.24) | 4.95 (4.60, 5.35) | <0.001 |
| **diabetes** |  |  |  | <0.001 |
| No | 16073091 (79%) | 13472888 (83%) | 2600203 (62%) |  |
| Yes | 4386124 (21%) | 2767515 (17%) | 1618609 (38%) |  |
| **eGFR** | 77.84 (21.87) | 79.55 (21.36) | 71.25 (22.59) | <0.001 |
| **Log NLR** | 0.74 (0.44, 1.03) | 0.72 (0.44, 1.01) | 0.79 (0.47, 1.12) | 0.085 |
| **smoker** |  |  |  | 0.002 |
| No | 9617396 (47%) | 7969079 (49%) | 1648317 (39%) |  |
| Yes | 10841819 (53%) | 8271324 (51%) | 2570495 (61%) |  |
| **Physical activity** |  |  |  | 0.033 |
| No | 11648250 (57%) | 9025336 (56%) | 2622914 (62%) |  |
| Yes | 8810965 (43%) | 7215067 (44%) | 1595898 (38%) |  |
| **depression** |  |  |  | <0.001 |
| No | 18052515 (88%) | 14615647 (90%) | 3436867 (81%) |  |
| Yes | 2406700 (12%) | 1624756 (10%) | 781945 (19%) |  |

Participants with CVD were significantly older than those without CVD (age: 66.83 vs. 59.56 years; p <0.001) and had a lower mean PIR (2.56 vs. 3.17; p<0.001). Approximately 60% of participants with CVD were female, and racial distributions differed significantly between the CVD and non-CVD groups (p=0.044).

The CVD cohort experienced a significantly higher burden of cardiometabolic and renal comorbidities. The prevalence of hypertension (87% vs. 69%; p<0.001) was significantly higher among those with CVD. Diabetes was significantly more prevalent among RA patients with concurrent CVD compared to those without cardiovascular complications (38% vs. 17%, p<0.001). Mean estimated glomerular filtration rate (eGFR) was significantly lower in the CVD group (71.25 vs. 79.55mL/min/1.73m²; p<0.001), indicating poorer baseline renal function. Lipid profiles demonstrated significantly lower HDL levels (51.57 vs. 56.07; p < 0.001), higher triglycerides (4.95 vs. 4.84; p < 0.001), and notably lower LDL cholesterol (97.94 vs. 113.02 mg/dL; p < 0.001) in the CVD group compared with the non-CVD group.

Behavioral and psychosocial risk factors were consistently more prevalent in the RA-CVD population. These individuals reported significantly higher rates of smoking (61% vs. 51%; p =0.002) and physical inactivity (62% vs. 56%; p =0.033), alongside a greater prevalence of moderate-to-severe depression (19% vs. 10%; p<0.001). BMI and NLR did not differ significantly between the CVD and non-CVD groups (p = 0.082 and p = 0.085) in RA population.

### Identification of Clinical Phenotypes

We identified six distinct data-driven phenotypes among participants with RA (Figure 2) by evaluating candidate solutions across k = 2–7. As shown in Figure 2, the six-cluster solution demonstrated favorable cluster cohesion, reproducible phenotype structure across independent train and test cohorts, and clinically interpretable separation patterns.

**Figure 2:**
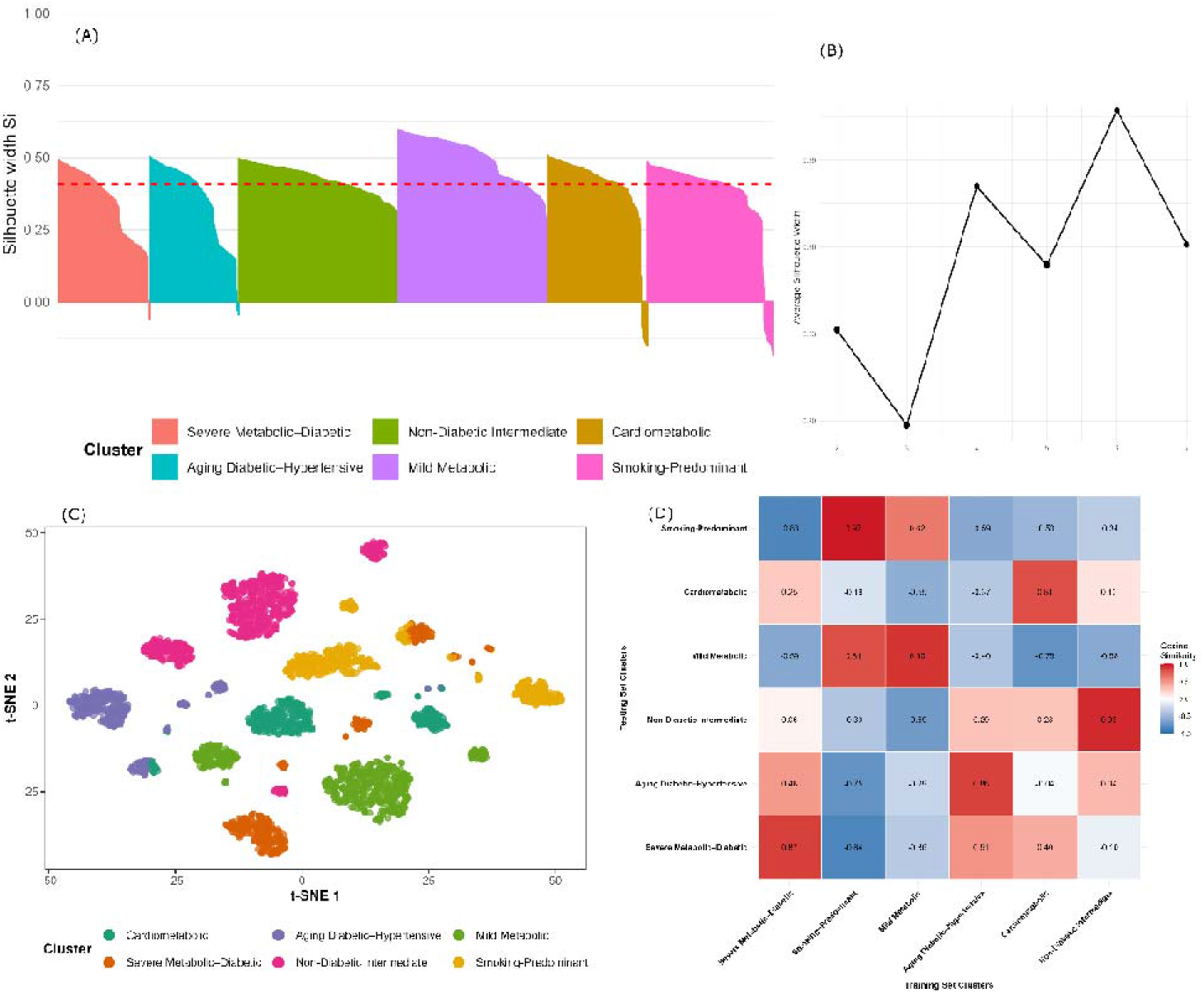
Validation, Optimization, Visualization, and Reproducibility of the Unsupervised Phenotypic Clustering Architecture. (A) Silhouette width plot for the optimal k=6 cluster solution. Individual bars represent the silhouette width (S_i_) for each participant. The red dashed line denotes the overall mean silhouette width. (B) Evaluation of clustering quality via average silhouette width across varying numbers of clusters (k), confirming the mathematical optimality of k = 6. (C) Visual distribution of the study population within a two-dimensional t-SNE embedding space, color-mapped by phenotypic cluster assignment to highlight distinct boundaries. (D) Cross-validation stability matrix display using Cosine Similarity coefficients between the training and testing set clusters.

The final model yielded an average silhouette width of 0.41 (Figure 2A), reflecting moderate but acceptable separation between clusters in a heterogeneous clinical population. Cluster sizes were well distributed, and no evidence of substantial cluster collapse or fragmentation was observed. Low-dimensional projection using t-SNE further demonstrated discernible grouping of individuals according to cluster membership (Figure 2C), supporting the structural validity of the identified phenotypes while acknowledging expected overlap inherent to complex cardiometabolic profiles.

Internal validation demonstrated reproducible recovery of phenotype signatures across datasets. Cosine similarity analysis showed strong correspondence between matched training and testing phenotypes, with coefficients ranging from 0.81 to 0.97 (mean = 0.89; Figure 2D). The Davies-Bouldin index showed a minor improvement from the training to the testing cohort (1.225 vs. 1.189), while the Dunn index remained virtually identical (0.025 vs. 0.029). The Calinski-Harabasz index reflected a change due to sample size scaling[26] (688.475 vs. 307.315) but maintained a highly significant variance ratio (see supplementary figure S3). Our findings support the internal reproducibility and consistency of the six-cluster solution, which was therefore retained for downstream analyses.

### Clinical Characterization of Cluster Phenotypes

Clinical heterogeneity was observed across the six phenotypes, which exhibited distinct combinations of cardiometabolic, inflammatory, and psychobehavioral characteristics (Figure 3).

**Figure 3:**
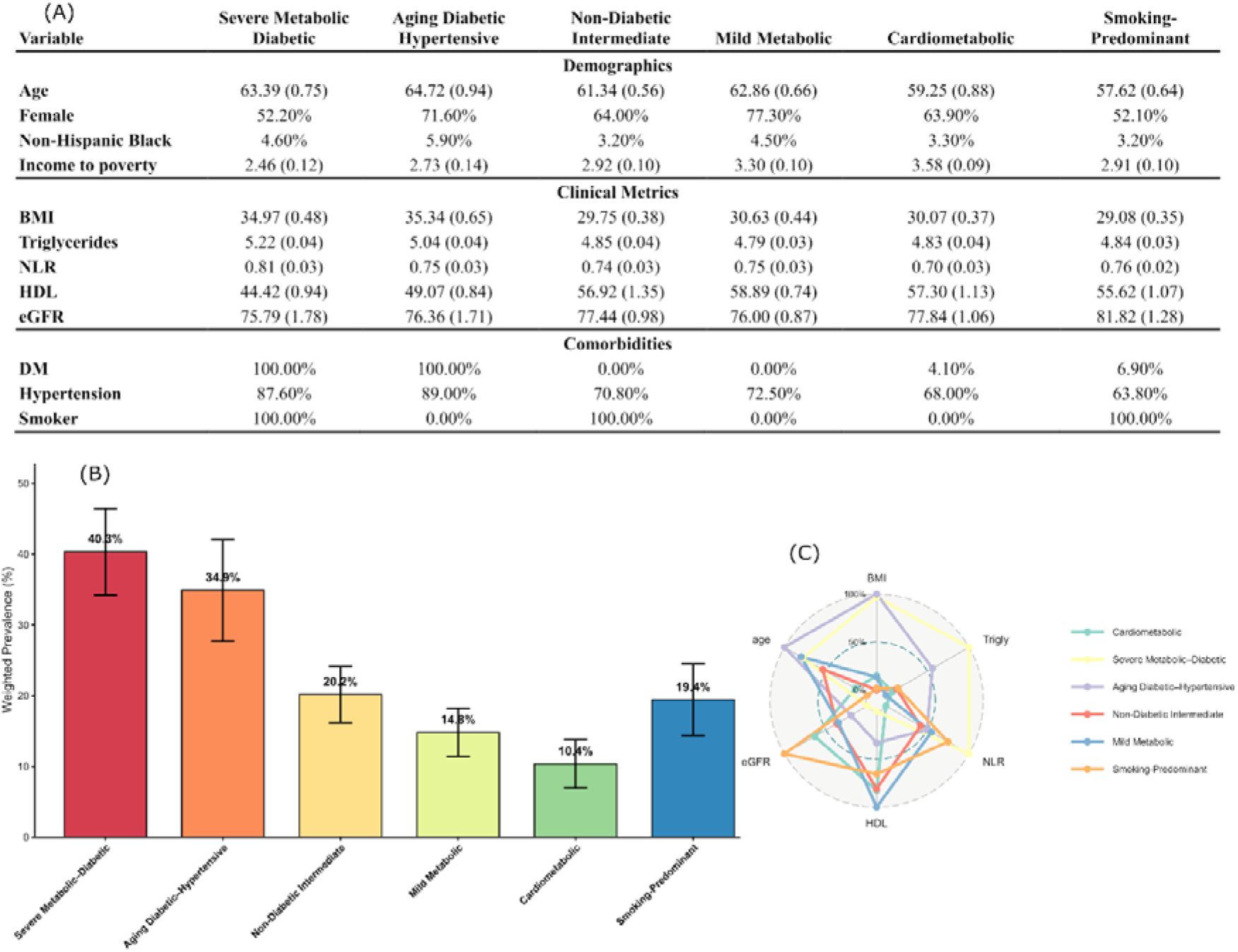
Cluster-specific phenotypic profiles and population-weighted prevalence of identified phenotypes. (A) Table displaying the distribution of key demographics across the six identified clusters. Continuous variables are presented as mean (SE), categorical variables are expressed as percentages (%). (B) Bar chart representing the population-weighted prevalence (%) with 95%CI for each cluster. (C) Normalized radar plot mapping the multi-dimensional clinical fingerprints and overlapping physiological shapes across continuous biometric domains, scaled on a relative percentage.

The Severe Metabolic–Diabetic phenotype exhibited the highest burden of adverse cardiometabolic features with elevated BMI, triglycerides, NLR, hypertension, and universal diabetes and smoking exposure (Figure 3A). This cluster also demonstrated the maximum 40.3% prevalence of CVD (Figure 3B). Additionally, radar plot mapped the overlapping physiological shapes across core continuous domains (Figure 3C).

The Aging Diabetic–Hypertensive phenotype had the oldest age distribution (64.72; SE=0.94), high prevalence of diabetes (100%) and hypertension (89%), reduced renal function (76.36; SE= 1.71), and comparatively higher NLR (0.75; SE = 0.03) and triglycerides (5.04; Se = 0.04). This phenotype demonstrated the second-highest burden of prevalent CVD (34.9%). In contrast, the Cardiometabolic phenotype had 10.4% CVD prevalence. It was characterized by comparatively higher eGFR (77.84; SE = 1.06), lower NLR, relatively preserved lipid parameters, and had 4.1% DM, 68% hypertension prevalence.

The Mild Metabolic phenotype cluster had 14.8% CVD burden. It was characterized by relatively preserved metabolic and inflammatory indices. In this group the PIR (3.30; SE= 0.10) was second highest and the HDL had highest level (58.89, SE=0.74) with no diabetic patients.

The Smoking-Predominant phenotype was uniquely characterized by universal smoking exposure (100% current smokers) despite a comparatively preserved metabolic profile, including a lower diabetes prevalence (6.9%) and relatively moderate BMI (mean BMI 29.08; SE = 0.35 kg/m²). Although systemic inflammatory burden was intermediate (NLR Z-score ≈ 0.6), this phenotype demonstrated a substantial weighted prevalence of CVD (19.4%) (see supplementary figure S5).

The Non-Diabetic Intermediate phenotype exhibited moderate cardiometabolic burden without overt diabetes mellitus (0%, Z-score ≈ -0.7), It had intermediate obesity (mean BMI 29.75 kg/m²; SE 0.38) and hypertension prevalence (70.8%). Despite the absence of diabetes, this phenotype demonstrated a CVD prevalence of 20.2%, illustrating that different combinations of cardiometabolic and behavioral characteristics were observed across phenotypes with varying CVD burden. Weighted prevalence estimates demonstrated a parallel gradient across phenotypes (Figure 3B). The Severe Metabolic–Diabetic and Aging Diabetic–Hypertensive phenotypes had the highest prevalence of CVD (40.3% and 34.9%, respectively), and the Cardiometabolic phenotype demonstrated the lowest prevalence (10.4%).

### Association Between Clinical Phenotypes and Cardiovascular Disease

Multivariable weighted binary logistic regression was used to evaluate associations between phenotype membership and prevalent CVD. The results are summarized in figure 4(A) and supplementary table S4. The cardiometabolic phenotype was considered as the reference category. Severe Metabolic–Diabetic phenotype exhibited the strongest association with prevalent CVD (aOR 4.11, 95% CI 2.56–6.59; *p* <0.001). Similarly, the Aging Diabetic– Hypertensive phenotype was associated with higher odds of prevalent CVD (aOR 3.38, 95% CI: 2.22–5.15; p < 0.001).

**Figure 4.**
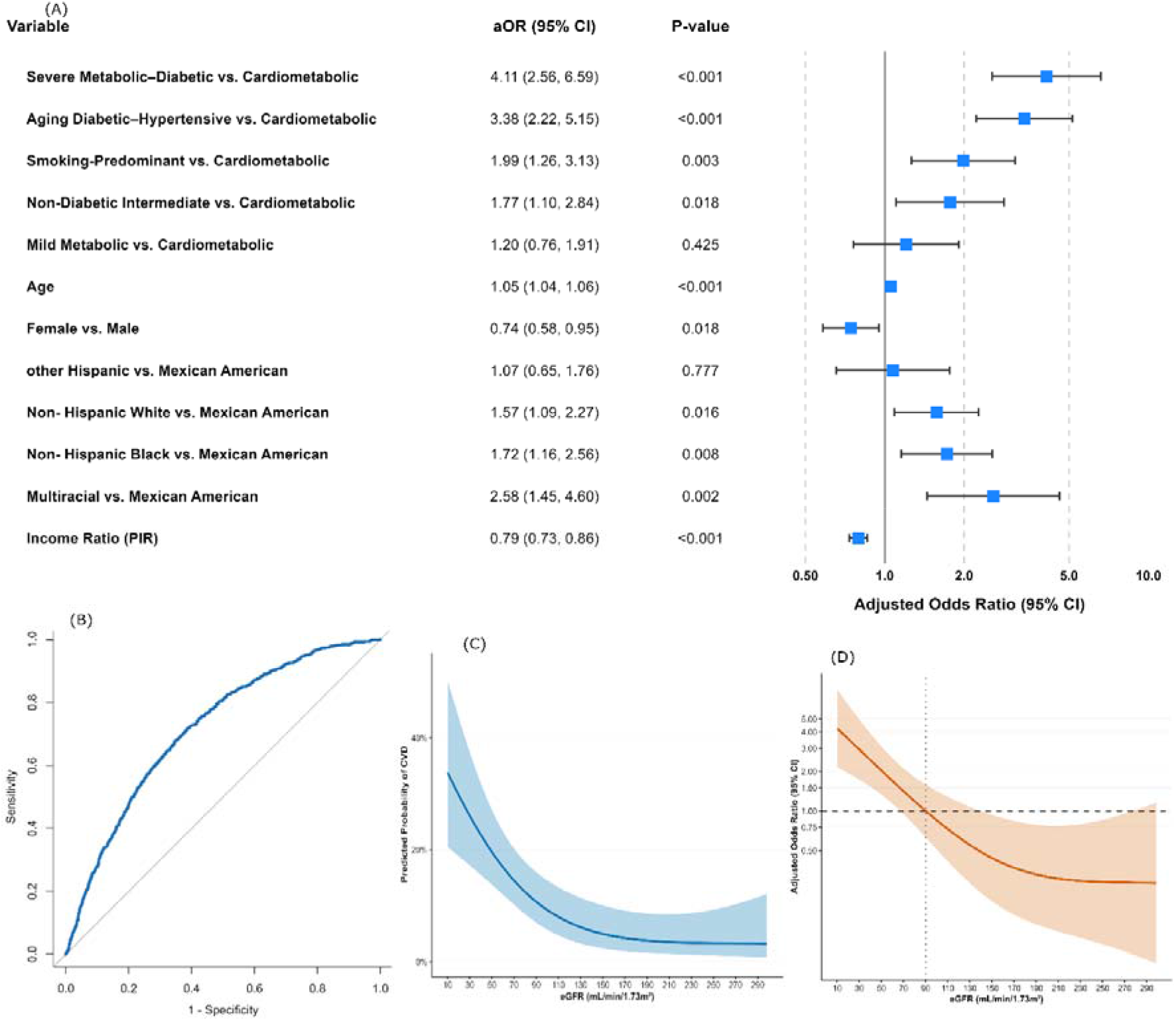
Association of cardiometabolic and psychobehavioral phenotypes with CVD and sensitivity analyses. (A) Survey-weighted adjusted odds ratios for prevalent CVD according to phenotype membership and sociodemographic covariates. (B) ROC curve demonstrating model discrimination. (C) Restricted cubic spline showing the estimated probability of prevalent CVD across eGFR. (D) Restricted cubic spline illustrating the nonlinear association between eGFR and prevalent CVD.

The Smoking-Predominant phenotype also demonstrated significantly higher odds of prevalent CVD (aOR 1.99, 95%CI 1.26–3.13; *p* = 0.003), whereas the Non-Diabetic Intermediate phenotype showed a modest but statistically significant association (aOR 1.77, 95%CI 1.10–2.84; *p* = 0.018). In contrast, the Mild Metabolic phenotype was not significantly associated with prevalent CVD after multivariable adjustment. Among covariates, older age was independently associated with higher odds of prevalent CVD (aOR 1.05, 95% CI 1.04–1.06; *p* <0.001), whereas higher PIR was associated with lower odds. Differences in the odds of prevalent CVD across racial groups were also observed.

The survey-weighted multivariable model demonstrated moderate discrimination for prevalent CVD, with an AUC of 0.718 (Figure 4B). As a sensitivity analysis, restricted cubic spline (RCS) modeling of eGFR demonstrated a significant nonlinear association with prevalent CVD (Figure 4C). Formal testing using a Wald χ² test provided significant evidence of nonlinearity (p = 0.005), indicating that the association between eGFR and prevalent CVD was not adequately represented by a simple linear term.

As shown in Figure 4C, the estimated probability of prevalent CVD increased progressively at lower levels of eGFR, with a steeper gradient below approximately 90 mL/min/1.73 m². Correspondingly, spline-based odds-ratio modeling showed higher adjusted odds of prevalent CVD at lower eGFR values, followed by attenuation and a plateau at higher levels of renal function (Figure 4D). Incorporation of nonlinear eGFR terms into the multivariable model did not materially alter the direction of the associations between phenotype membership and prevalent CVD (See supplementary table S4). It supports the robustness of the identified multidimensional phenotypes independent of isolated renal dysfunction.

## Discussion

Our study identified six distinct cardiometabolic and psychobehavioral phenotypes among adults with RA, with substantial heterogeneity in CVD burden across groups. These data-driven phenotypes should not be interpreted as fixed biological subtypes, but rather as clinically interpretable patterns representing a continuum of multidimensional cardiovascular risk. The highest-risk profiles were characterized by combinations of metabolic dysfunction, diabetes, hypertension, renal impairment, systemic inflammatory burden, and adverse behavioral characteristics. These findings are consistent with evidence that cardiovascular risk in RA is heterogeneous and is not fully captured by individual risk factors or conventional cardiovascular risk algorithms[5], [17].

The identification of heterogeneous profiles is consistent with previous clustering studies in RA, although the clinical meaning of the clusters has differed according to the variables and outcomes examined. For example, Curtis et al. identified five RA subgroups characterized by differences in disease activity, multimorbidity, metabolic and psychiatric comorbidity, and disease duration[27]. Similarly another population-based study identified recurring comorbidity patterns in RA, including cardiovascular and mental/behavioral profiles[28]. A more recent clustering studies have identified distinct groups according to combinations of comorbidity burden and RA-related characteristics[29]. In contrast to these approaches, which primarily characterized RA disease heterogeneity or overall comorbidity burden, our study focused specifically on CVD heterogeneity and jointly incorporated metabolic, inflammatory, renal, behavioral, and psychosocial characteristics. Thus, our findings extend previous RA clustering research by identifying multidimensional cardiovascular-risk profiles rather than general RA or comorbidity subgroups.

The Severe Metabolic–Diabetic and Aging Diabetic–Hypertensive phenotypes had the highest observed CVD burden. The Severe Metabolic–Diabetic phenotype was characterized by high BMI, hypertriglyceridemia, elevated NLR, diabetes, and smoking, whereas the Aging Diabetic–Hypertensive phenotype combined older age, diabetes, substantial hypertension, and reduced renal function. These profiles are consistent with established evidence linking diabetes, metabolic dysfunction, hypertension, inflammation, and renal impairment to cardiovascular vulnerability in RA[6], [30], [31]. Importantly, the clustering framework illustrates how this risk characteristics co-occurred within distinct patient profiles rather than acting as isolated risk factors. The nonlinear association between eGFR and prevalent CVD, with the estimated probability of prevalent CVD increasing more sharply below approximately 90 mL/min/1.73 m², further supports the relevance of renal function within this multidimensional risk structure[32]. The clustering framework therefore captured combinations of risk characteristics that were associated with different levels of CVD burden.

The Smoking-Predominant phenotype provided a distinct behavioral pattern within the cohort. This group was characterized by predominant smoking exposure and showed elevated CVD burden with a relatively preserved metabolic profile. This finding is consistent with evidence linking smoking with cardiovascular and inflammatory risk in RA [30] [33], and illustrates why behavioral characteristics may complement conventional metabolic risk assessment. Smoking remains an important modifiable cardiovascular risk factor in RA and is not always adequately addressed in routine rheumatologic care [34] [35]. The Non-Diabetic Intermediate phenotype showed moderate obesity and hypertension without diabetes, highlighting an intermediate cardiometabolic profile [36],[37]. The Mild Metabolic phenotype had relatively preserved metabolic characteristics, including higher HDL and absence of diabetes, and showed a CVD prevalence of 14.8% with a non-significant adjusted association. Together, these profiles illustrate that cardiovascular-risk heterogeneity in RA may arise from different combinations of metabolic and behavioral characteristics rather than from a single dominant risk factor.

The Cardiometabolic phenotype was used as the reference category because it had the lowest CVD prevalence. Its relatively low NLR and preserved eGFR were consistent with a comparatively favorable CVR profile [38], while previous evidence supports the relevance of inflammatory biomarkers for CVR beyond their role in disease activity assessment [39]. The Mild Metabolic phenotype showed intermediate obesity and metabolic abnormalities, its association with CVD was attenuated after adjustment, suggesting that its relatively preserved HDL and lower inflammatory burden may characterize a comparatively lower-risk profile.

The six-cluster solution also demonstrated internal reproducibility across the independent training and testing cohorts. Corresponding clusters showed high cosine similarity, while the Davies–Bouldin and Dunn indices were broadly comparable between partitions. The average silhouette width of 0.41 indicated moderate cluster separation, supporting the internal consistency of the clustering solution[40]. These findings support the reproducibility of the identified data driven profiles across independent subsets of the study population, although they should not be interpreted as fixed biological entities.

In this study, we found several sociodemographic characteristics were associated with prevalent CVD. Older age was strongly associated with higher CVD odds, whereas female sex was associated with lower odds than male sex, consistent with previous evidence of greater cardiovascular morbidity and faster atherosclerotic progression among men with RA [41]. Differences in CVD odds across racial and ethnic groups were also observed, consistent with evidence that cardiovascular outcomes in RA may reflect both biological factors and broader social and structural determinants [42]. Higher PIR was associated with lower CVD odds, potentially highlighting the relationship between socioeconomic circumstances, access to care, health literacy, and preventive health behaviors[43]. These associations provide additional context for cardiovascular heterogeneity within the RA population.

An important consideration is that the present clustering framework was developed entirely within the RA population and did not incorporate RA-specific measures such as disease activity, serologic status, disease duration, or disease-modifying treatment exposure. Therefore, RA specificity cannot be established. The identified profiles should be interpreted as multidimensional cardiovascular-risk patterns observed among individuals with RA rather than as distinct RA-specific biological subtypes. Comparative studies incorporating non-RA populations and detailed RA-specific clinical measures are needed to determine whether these profiles are unique to, enriched in, or modified by RA.

These findings have potential implications for cardiovascular risk assessment in RA. Current EULAR recommendations apply a general RA-related risk adjustment despite substantial heterogeneity in cardiometabolic and other risk characteristics [16]. The present results suggest that phenotype-informed assessment could complement conventional risk evaluation by identifying patients with different combinations of metabolic, renal, inflammatory, and behavioral characteristics. In particular, the Smoking-Predominant profile indicates that behavioral risk may remain clinically relevant even in the absence of severe metabolic abnormalities. Depression and physical activity were also incorporated into the clustering framework, extending cardiovascular-risk characterization beyond conventional metabolic measures [44], [45]. Because the variables used in the clustering framework are generally obtainable through routine clinical assessment and laboratory testing [46], this approach may be feasible for further evaluation in clinical settings.

The findings support consideration of multidimensional cardiovascular-risk profiles alongside conventional risk assessment in RA. However, the clinical utility of these profiles for treatment selection or individualized prevention remains to be established. Prospective studies are needed to determine whether these profiles are associated with incident cardiovascular events and whether phenotype-informed approaches provide clinically meaningful improvement in cardiovascular risk assessment.

## Conclusion

This study identified substantial heterogeneity in cardiovascular-risk profiles among individuals with RA that may not be fully captured by conventional single-risk-factor approaches. Using a data-driven phenotyping framework among adults with RA, we identified six distinct cardiometabolic and psychobehavioral phenotypes that exhibited substantial differences in CVD burden and adjusted odds of prevalent CVD. The profiles with the highest observed CVD burden were characterized by combinations of metabolic dysfunction, systemic inflammation, renal impairment, and adverse behavioral characteristics, whereas more metabolically preserved phenotypes exhibited comparatively lower CVD burden. Notably, smoking characterized a distinct phenotype with elevated CVD burden despite relatively preserved metabolic characteristics, while depression and physical activity were incorporated as behavioral and psychosocial dimensions of the observed cardiovascular-risk profiles. These findings highlight the importance of considering multidimensional risk profiles, including behavioral and psychosocial characteristics, when assessing cardiovascular vulnerability in RA. Our findings support further evaluation of phenotype-based approaches to multidimensional CVR assessment in RA, although their clinical utility for prevention and management remains to be established. However, these phenotypes should not be interpreted as RA-specific biological subtypes, as RA disease activity, serologic status, and treatment exposure were not incorporated into the clustering framework. Future longitudinal studies incorporating RA disease activity, serologic status, treatment exposure, and incident cardiovascular outcomes are needed to validate the prognostic and clinical utility of these phenotypes.

### Limitations of the study

Several limitations should also be considered. First, the cross-sectional design precludes causal inference, and temporal relationships between phenotype membership and CVD cannot be established. Second, RA was identified using self-reported physician diagnosis rather than validated ACR/EULAR classification criteria. Consequently, some participants may have had osteoarthritis, gout, or other forms of arthritis, introducing potential disease misclassification that could influence both phenotype assignment and the estimated associations with CVD. Third, NHANES does not provide standardized RA-specific measures of disease activity (e.g., DAS28), disease duration, anti-CCP antibodies, ESR, or information on synthetic and biologic DMARD exposure that could be consistently incorporated into the present analysis. Consequently, associations between RA-specific disease activity, serologic characteristics, disease duration, treatment exposure, and the observed cardiovascular-risk profiles could not be directly evaluated. In addition, pharmacotherapy information regarding glucocorticoid exposure, was not incorporated into the present analysis. Glucocorticoid therapy may contribute to hyperglycemia and treatment-associated or secondary diabetes, complicating the distinction between underlying diabetes and treatment-related dysglycemia. Although HbA1c ≥6.5% was used together with self-reported clinical diagnosis to identify diabetes and has the advantage of not requiring fasting, medication-related effects could not be fully accounted for and may introduce residual uncertainty in diabetes classification and phenotype assignment. Residual confounding from unmeasured variables also remains possible. Finally, the composite CVD outcome combined myocardial infarction, angina, heart failure, and stroke into a single endpoint. These conditions may have distinct pathophysiological mechanisms and may differ in their associations with the identified CVR profiles; therefore, the present analysis does not establish whether the observed phenotype–CVD associations are consistent across individual cardiovascular outcomes. External validation in prospective and non-US RA populations will be necessary to determine the generalizability of the identified phenotypes.

**Figure S1.** Missing Data Profiling and Multiple Imputation Diagnostics via Chained Equations (MICE). **Table S1.** Unweighted Baseline Demographics, Clinical, and Psychobehavioral Characteristics of the Study Sample Stratified by Cardiovascular Disease Status. **Figure S2**. Validation of Clustering Framework via Side-by-Side Average Silhouette Width Across Train and Test Partitions. **Figure S3.** Comparison of Cluster Quality Indices Between Training and Testing Dataset Partitions. **Figure S4.** t-SNE Projection Mapping of Derived Phenotypes Across Training and testing Cohort Splits. **Figure S5.** Stratum-Specific Z-Score Heatmap Biplots Across Internal Cross-Validation Sets. **Table S2.** Multicollinearity Diagnostics via Variance Inflation Factors (VIF) for the Primary Regression Framework. **Table S3.** Weighted Multivariable Binary Logistic Regression Evaluating the Association Between Phenotypic Clusters and Prevalent Cardiovascular Disease. **Table S4.** Sensitivity Analysis via Population-Weighted Multivariable Logistic Regression Adjusting for Non-Linear Renal Function Using Restricted Cubic Splines (RCS). **Figure S6.** Distributions of triglycerides and neutrophil-to-lymphocyte ratio before and after log transformation.

## Data Availability

The dataset(s) supporting the conclusions of this article are available in the National Health and Nutrition Examination Survey (NHANES) repository maintained by the National Center for Health Statistics (NCHS), Centers for Disease Control and Prevention (CDC), at https://wwwn.cdc.gov/nchs/nhanes/continuousnhanes/default.aspx. All data analysed during the current study are publicly available and can be accessed without restriction.

## Abbreviations

CVD: Cardiovascular disease
CVR: Cardiovascular Risk
RA: rheumatoid arthritis
NHANES: National Health and Nutrition Examination Survey
PAM: Partitioning Around Medoid
t-SNE: t-distributed stochastic neighbor embedding
MICE: multiple imputation by chained equations
RCS: restricted cubic spline
eGFR: estimated glomerular filtration rate
EULAR: European League Against Rheumatism
MEC: Mobile Examination Center
PHQ: Patient Health Questionnaire
aOR: adjusted odds ratio
AUC: Area Under Curve
PIR: Poverty-Income Ratio
LDL: Low density lipoprotein
HDL: High density lipoprotein
NLR: Neutrophil-to-Lymphocyte Ratio
MDRD: Modification of Diet in Renal
RCS: Restricted Cubic Splines

## Ethics approval and consent to participate

NHANES is a nationally representative research program conducted by the Centers for Disease Control and Prevention (CDC). The NHANES protocol was approved by the National Center for Health Statistics (NCHS) Research Ethics Review Board, and all participants provided written informed consent prior to participation. Because NHANES data are publicly available, fully de-identified, and accessible through the CDC website, no additional institutional review board approval or participant consent was required for the present secondary data analysis.

## Authors Contribution

S.T.J and M.M.H were responsible for the study design, S.T.J provided statistical support, drafted the manuscript, and were primarily responsible for the final content. S.T.J., A.A., S.H., were involved in data curation, and preparation, S.T.J, S.H., A.A. analyzed the data and the visualization of the results. M.S.R, M.M.A., M.M.R., M.M.H. validated the statistical analysis, edited, reviewed the manuscript. All authors actively participated in the research process made substantial contributions to manuscript and approved the final version.

## Acknowledgements

We would like to acknowledge all patients and involved health personnel.

